# Dengue epidemic alert thresholds, a tool for surveillance and epidemic detection

**DOI:** 10.1101/2024.10.22.24315684

**Authors:** Maile B. Thayer, Melissa Marzan-Rodriguez DrPH, Jomil Torres Aponte, Aidsa Rivera DrPH, Dania M. Rodriguez, Zachary J. Madewell, Kristyna Rysava, Gabriela Paz-Bailey, Laura E. Adams, Michael A. Johansson

**Affiliations:** Centers for Disease Control and Prevention, San Juan, Puerto Rico; Puerto Rico Department of Health, San Juan, Puerto Rico

**Keywords:** dengue, dengue virus, epidemics, epidemic thresholds, epidemic detection

## Abstract

Epidemic detection enables swift public health responses. Dengue viruses pose a significant public health challenge in Puerto Rico, where they are endemic and cause intermittent epidemics. A weekly intercept-only negative binomial regression model was fitted using historical data from January 1986 to June 2024. Thresholds were defined using three percentiles (60%, 75%, and 90%). The 75th percentile threshold aligned best with historical epidemic classifications. This model provides a robust method for defining thresholds, accounting for skewed data, utilizing all historical data, and improving upon traditional methods like endemic channels. In March 2024, the Puerto Rico Department of Health declared a public health emergency due to an unseasonably early surge in cases that exceeded the epidemic alert threshold in February. This real-time application highlights the value of these thresholds to support dengue epidemic detection and public health response. Integrating thresholds with other tools and strategies can enhance epidemic preparedness and management.

**One-sentence summary line:** Epidemic alert thresholds can correctly detect and classify epidemics and enable timely public health response.

## Introduction

Epidemics can have rapid and major impacts on population health and health systems. While predicting epidemics remains challenging, timely detection enables public health officials to respond swiftly and effectively. In endemic areas, where disease is consistently present, epidemic warnings must be carefully developed and linked to actionable responses to optimize public health efforts. In places where diseases are endemic, it can be difficult to distinguish baseline levels from epidemic activity due to the constant presence of the disease. There is also the potential for underestimation of epidemic severity due to adaptation and partial immunity within the population, fluctuations in case reporting and possible changes in diagnostic practices. In non-endemic areas, developing accurate epidemic detection mechanisms is often more complex due to low baseline levels of the disease. The U.S. Centers for Disease Control and Prevention (CDC) defines an epidemic as “an increase, often sudden, in the number of cases of a disease above what is normally expected in that population in that area” (1). For a more precise definition tailored to specific diseases, regions, and timeframes, factors such as frequency, duration, amplitude, and overall burden of cases should be considered. These elements are essential for defining disease-and region-specific epidemic alert thresholds— levels of disease incidence that trigger a necessary response. These definitions must try to strike a balance between minimizing false alarms (too many epidemic alerts) and failures to correctly identify an epidemic (not enough epidemic alerts). These elements are crucial for defining epidemic alert thresholds, the number of cases that must be exceeded to signal an alarm for an epidemic.

Dengue viruses (DENV) impose a considerable economic and health burden to the areas affected, particularly during epidemic years. DENV are mosquito-borne viruses that present an increasing public health challenge in tropical and subtropical regions (2, 3). Roughly half of the global population currently lives in areas suitable for DENV transmission, with the majority of those affected residing in Asia, followed by Africa and the Americas (4).

Puerto Rico, a United States territory in the Caribbean, has been greatly affected by DENV in recent decades and accounts for over 95% of locally acquired cases in the United States. Transmission is endemic and seasonal, with large epidemics occurring roughly every four to five years (5). A large DENV epidemic in Puerto Rico took place in 2012–2013, resulting in over 18,000 suspected and 9,200 confirmed cases (6–8). This period was followed by several years of relatively lower case counts until March 2024, when a concerning surge prompted a public health emergency declaration by Puerto Rico’s Department of Health due to an unseasonably early increase in cases.

States and policymakers define dengue epidemic alert thresholds in endemic regions using different methods, with varying results. One common approach, known as “endemic channels,” establishes thresholds by analyzing recent data (often excluding “epidemic years”) using a measure of yearly variability (usually standard deviation) and a measure of central tendency (usually the mean) to set an upper bound of expected case numbers (e.g., 2 standard deviations above the mean) (9–12). Although widely used, this method has limitations, particularly the assumption that data (reported case counts of dengue) are normally distributed. In reality, epidemiological data are usually skewed, with many low counts and few very high counts. Assuming that the data are normally distributed therefore results in thresholds that do not accurately reflect the actual distribution of cases. This approach may even produce impossible values, such as the expectation of negative case counts in low transmission years. Most existing dengue threshold definitions depend on this assumption of normality, using variations of the mean as a measure of central tendency (10, 13). Similarly, endemic channel threshold implementations often consider only the last several (usually 5-8) years of data, excluding epidemic years or years with low case counts, such as during the COVID-19 pandemic (9, 10). This not only omits some available data, but also overlooks valuable information from epidemics—the very events the thresholds are designed to detect.

Public health officials from the CDC and the Puerto Rico Department of Health (PRDH) have closely collaborated to develop and implement an improved method for defining dengue epidemic alert thresholds. This approach, currently used for classifying and detecting epidemics in Puerto Rico, allows for data heterogeneity and makes full use of historical data. This tool is currently used as a part of DENV surveillance, integrated in weekly epidemic assessments, and featured in the PRDH’s weekly arboviral disease reports.

## Methods

### Data

This thresholds analysis utilizes historical dengue surveillance data from Puerto Rico’s Passive Arboviral Disease Surveillance System, including all probable and confirmed DENV cases (including private laboratory results) reported by healthcare providers from January 1, 1986-June 30, 2024. Suspected dengue cases are reported based on clinical suspicion to the surveillance system, jointly managed by PRDH and CDC, with laboratory confirmation in PRDH. Probable cases are defined as those with a positive immunoglobulin M result for DENV, and confirmed cases are defined as those with a positive reverse transcription polymerase chain reaction result for DENV. Cases are aggregated on a weekly timescale.

### Statistical Analysis

*Negative binomial model*. We calculate weekly dengue alert thresholds using retrospective historical data by fitting a negative binomial intercept-only regression model for each week. This model incorporates all prior data points from the same week in previous years. The negative binomial distribution models count data (in this case, dengue case count data) while allowing for a flexible relationship between the mean and variance and can account for skewed data, as is often seen with dengue case counts. Unlike traditional methods which assume normally distributed case counts, the negative binomial distribution effectively accounts for epidemic years and uses the full dataset. Note that a negative binomial intercept-only regression model is equivalent to a negative binomial distribution; however, the formulation of the model in a regression framework allows for possible variations with predictors and lagged terms.

The negative binomial regression model allows us to extract quantile values that serve as dengue alert thresholds for comparison with current weekly case counts. For instance, the 50th percentile represents the historical median, which is not the actual median from historical counts but rather the median estimated by the model based on all historical data. This approach provides an estimate of the “typical” case count. Higher quantiles provide insights into epidemics, which are not expected annually. We assess three percentiles for epidemic detection: 60%, 75%, and 90%. These percentiles roughly correspond to expected epidemics occurring once every 2.5, 4, and 10 years, respectively 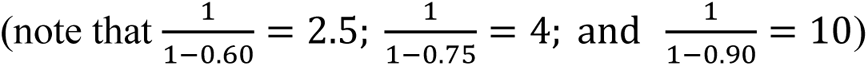. Estimated thresholds are based on data for previous years and can be updated annually as new data become available. Each week’s threshold is derived from its corresponding historical week. As data accumulates over time, the thresholds become progressively more stable.

After calculating the thresholds using the negative binomial regression model, we smooth the threshold trajectories to eliminate noise and random variation in the week-to-week threshold patterns. We apply centered linear filtering with convolution using a fast Fourier transform over 11 weeks (including the week of interest, plus 5 weeks before and 5 after each data point). This technique calculates a moving average of thresholds across the chosen window of 11 weeks. Centered linear filtering assigns weights to the data points within the window, with the current week receiving the highest weight and weights gradually decreasing for weeks further away. This approach minimizes the influence of distant weeks while preserving overall trends. The fast Fourier transform efficiently performs the convolution, making it faster than calculating the weighted average directly. The 11-week window was chosen after performing a sensitivity analysis across a range of windows and selecting the one with the best balance between removing noise removal and maintaining the main signal in the data (Fig S2). For windows less than 11, there were still some fluctuations due to noise, and for windows greater than 11, there was not much improvement on capturing the overall signal.

All analyses were performed using R version 4.4.0 (14). We utilized the ’MASS’ package for fitting the negative binomial model (function ’glm.nb’) and package ’stats’ for smoothing (function ’filter’) (15, 16). Code with example data is available on GitHub: https://github.com/maile-thayer/epidemic_thresholds.

## Results

From January 1986 to June 2024 a total of 86,282 confirmed or probable DENV cases were reported, with an annual mean of 2,212 cases (median: 1,533; range: 40 to 10,356). Histograms of yearly cases demonstrate right skewness, with more pronounced skewness in recent data due to more frequent low case counts since 2013 (Figure 1, Figure S1). Cases typically peaked around October and reached a yearly low in April. Annually fitted thresholds from 2001 showed general stability over time, with slight increases following the 2010 and 2012–2013 epidemics, and a gradual decrease during subsequent low transmission years (Figure 2).

**Figure 1.**
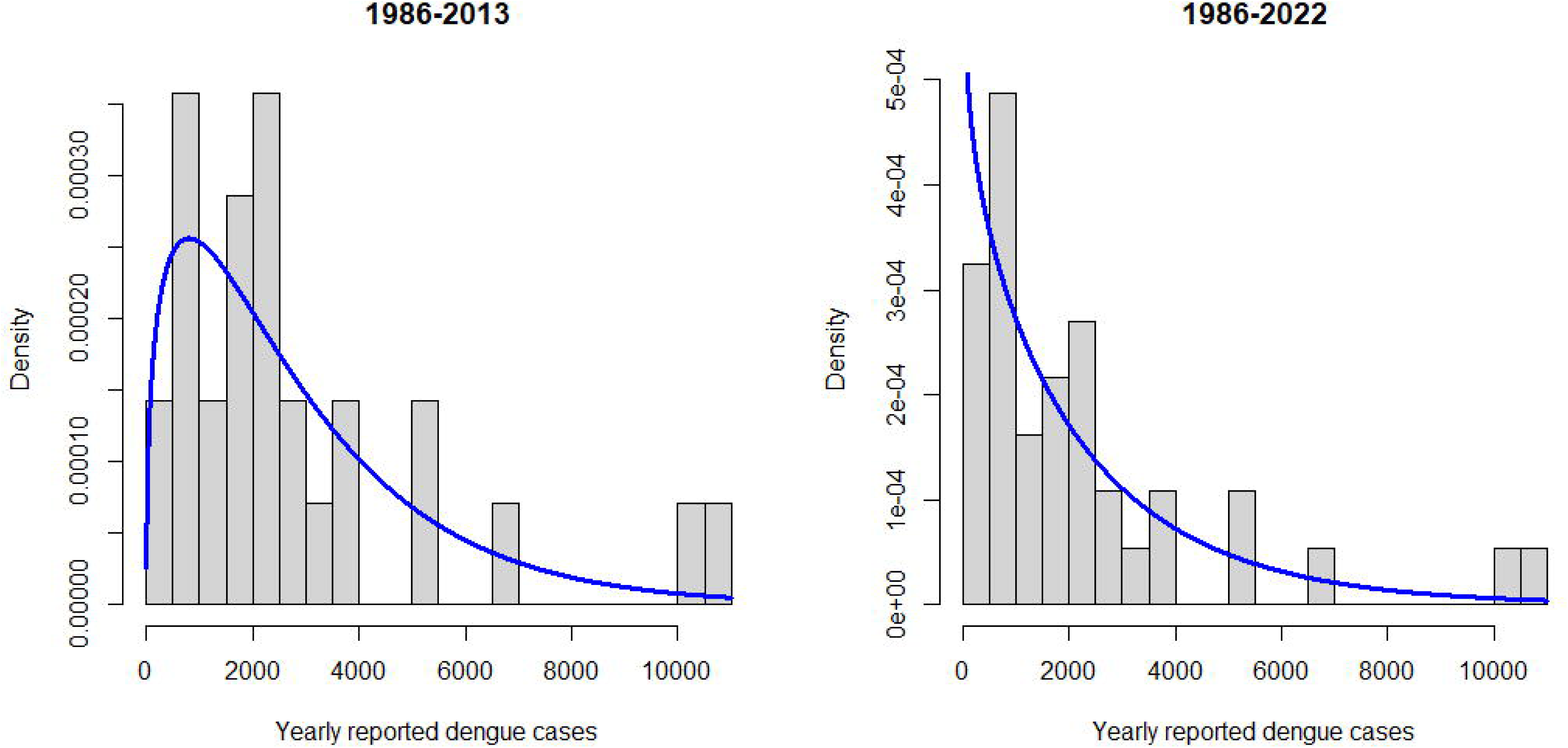
Histograms of total yearly cases from 1986–2013 and 1986–2023 in Puerto Rico, fit with negative binomial distributions. The densities of annual reported probable and confirmed island-wide dengue cases are shown (grey bars), with the corresponding fitted negative binomial distribution in blue. On the left are pre-Zika and chikungunya data (1986– 2013) and on the right are all available historical data as of January 1, 2024 (1986–2023).

**Figure 2.**
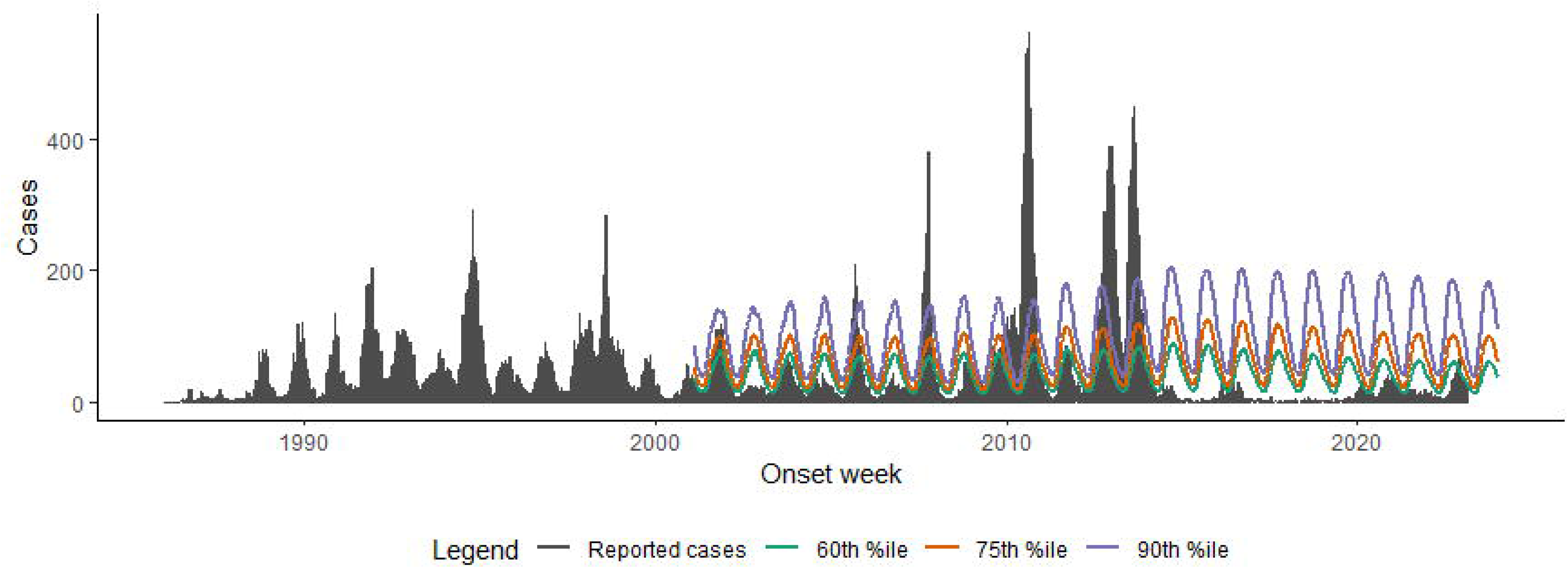
Reported probable and confirmed dengue virus cases and three estimated outbreak thresholds – Puerto Rico, 1986–2024. The weekly reported probable and confirmed dengue cases (≥1 positive PCR or IgM result) are represented by the grey bars. Weekly thresholds were calculated at the beginning of each new year starting in 2001 and are represented by the colored lines (60^th^ percentile in green; 75^th^ percentile in orange; and 90^th^ percentile in purple).

To evaluate the performance of the proposed thresholds in identifying epidemic years, we compared them with data from 2001–2023. The largest epidemics during this period occurred in 2007, 2009, 2010, 2012, and 2013. The three threshold definitions mostly agreed on identifying epidemic years, detecting epidemics in 2001, 2005, 2007, 2010, 2012, and 2013. However, there were discrepancies, resulting in variations in numbers of identified outbreaks and different timing of when the epidemics would have been detected (Figure 3). With a threshold at the 60^th^ percentile, the alert threshold would have been exceeded in 14 of the 23 years, including 6 years that would not have been detected using higher threshold definitions. In 2009, cases exceeded the 60^th^ percentile threshold and then dropped below it six different times. In 2020, cases exceeded the 60^th^ percentile two separate times. With a threshold at the 90% percentile, the threshold would have been exceeded in only six of the 24 years. In two of those six years identified, cases exceeded and then dropped below the 90^th^ percentile threshold twice in 2010 and then 16 times in 2012. Comparing these thresholds with alternative classifications revealed that 60% was too low, detecting epidemics in years with manageable case numbers, yet 90% was too high; for example, using 90% missed 2009 when case numbers were high and exceeded and then dropped below the threshold multiple times in 2010 and 2012. After consultation with PRDH, the 75^th^ percentile was found to align best with their experience and management response required for previous epidemics.

**Figure 3.**
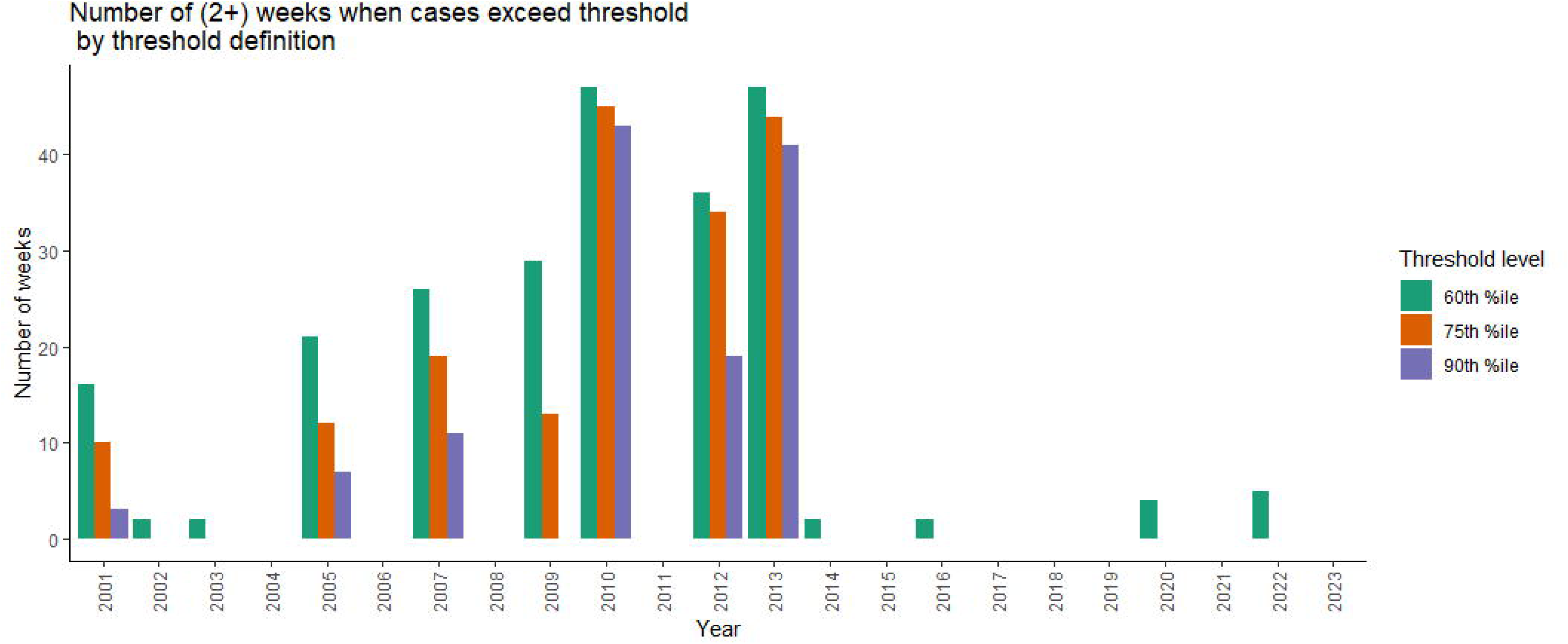
Number of weeks (2 or more) per year when dengue cases exceed threshold, with differing definitions: Puerto Rico January 2001 - June 2024. The number of weeks when the reported probable or confirmed cases are above the dengue threshold are tallied for each year using different threshold definitions, represented by the different-colored bars. A threshold using the 60^th^ percentile is shown in green, the 75^th^ percentile (currently used) is shown in orange, and the 90^th^ percentile is shown in purple. Two or more consecutive weeks above the threshold is considered an epidemic. Note that 2024 is shown, but data are only included up to the end of June.

An epidemic is then considered by PRDH as two or more consecutive weeks when reported probable and confirmed cases exceed the weekly (75^th^ percentile) epidemic alert threshold. Using the 75^th^ percentile alert threshold definition along with a requirement that reported probable and confirmed cases exceed this threshold for at least two consecutive weeks, there were six years with outbreaks detected, with long periods above the threshold occurring in 2010 and 2013 (45 weeks each, Figure 3). This threshold definition accurately identifies the six years (2001, 2005, 2007, 2010, 2012, and 2013) that PRDH identified as epidemic years. The outbreaks in 2010 and 2013 were also the years with the most cases above the thresholds (i.e., 8,273 and 7,011 excess cases, respectively). Weekly alert thresholds ranged from a low of 18 cases during the low season in spring to a high of 126 during peak transmission months in fall (mean: 62; median: 59) from 2001– June 2024. For other settings, it is important to collaborate with local health departments and policy makers to define the threshold level and decide upon the number of consecutive weeks needed to define epidemics.

### Two real-time uses of thresholds: 2022 and 2024 dengue in Puerto Rico

At the end of 2022, there was a rapid increase of DENV cases in Puerto Rico. Although the case numbers were higher than usual (exceeding the historical median) starting in October 2022, they approached but never surpassed the epidemic alert threshold (Figure 4A). With the weekly thresholds available, PRDH and the CDC monitored the status of the case counts each week and concluded that while cases were above the historical median, they never exceeded the thresholds and hence did not trigger an epidemic alert. Additionally, the historical median and threshold helped predict that dengue case numbers would decrease at the beginning of the year, which was accurately reflected in the downturn in early 2023.

**Figure 4.**
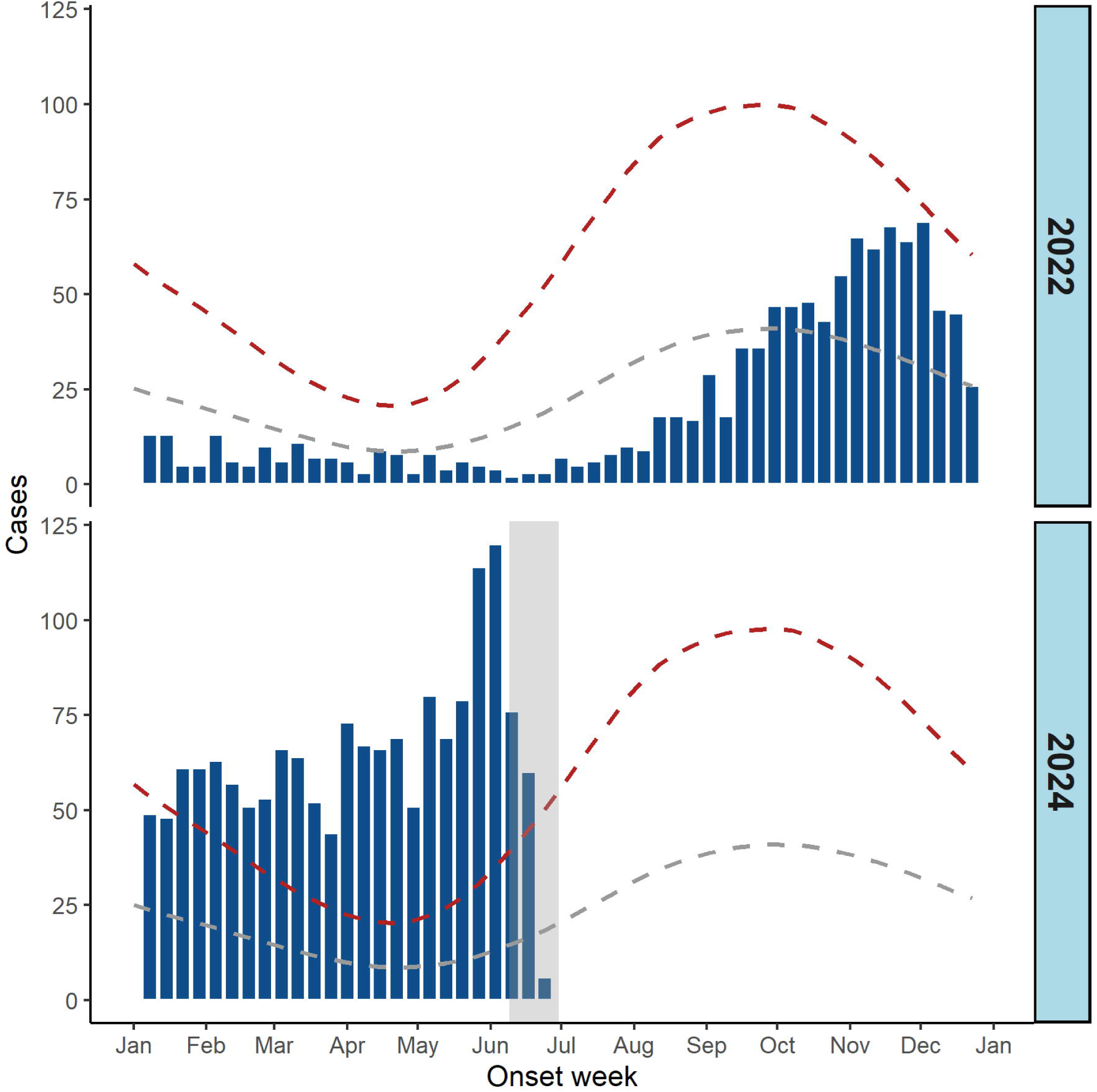
Reported dengue cases compared to historical weekly dengue alert thresholds in Puerto Rico, 2022 and 2024. The weekly reported probable and confirmed cases (≥1 positive PCR or IgM result) are represented by the navy bars, while the weekly historical median and threshold are represented by the dashed red and grey lines, respectively. The last three weeks of data in 2024 are highlighted in the light grey rectangle to denote that the most recent weeks are likely subject to reporting delays.

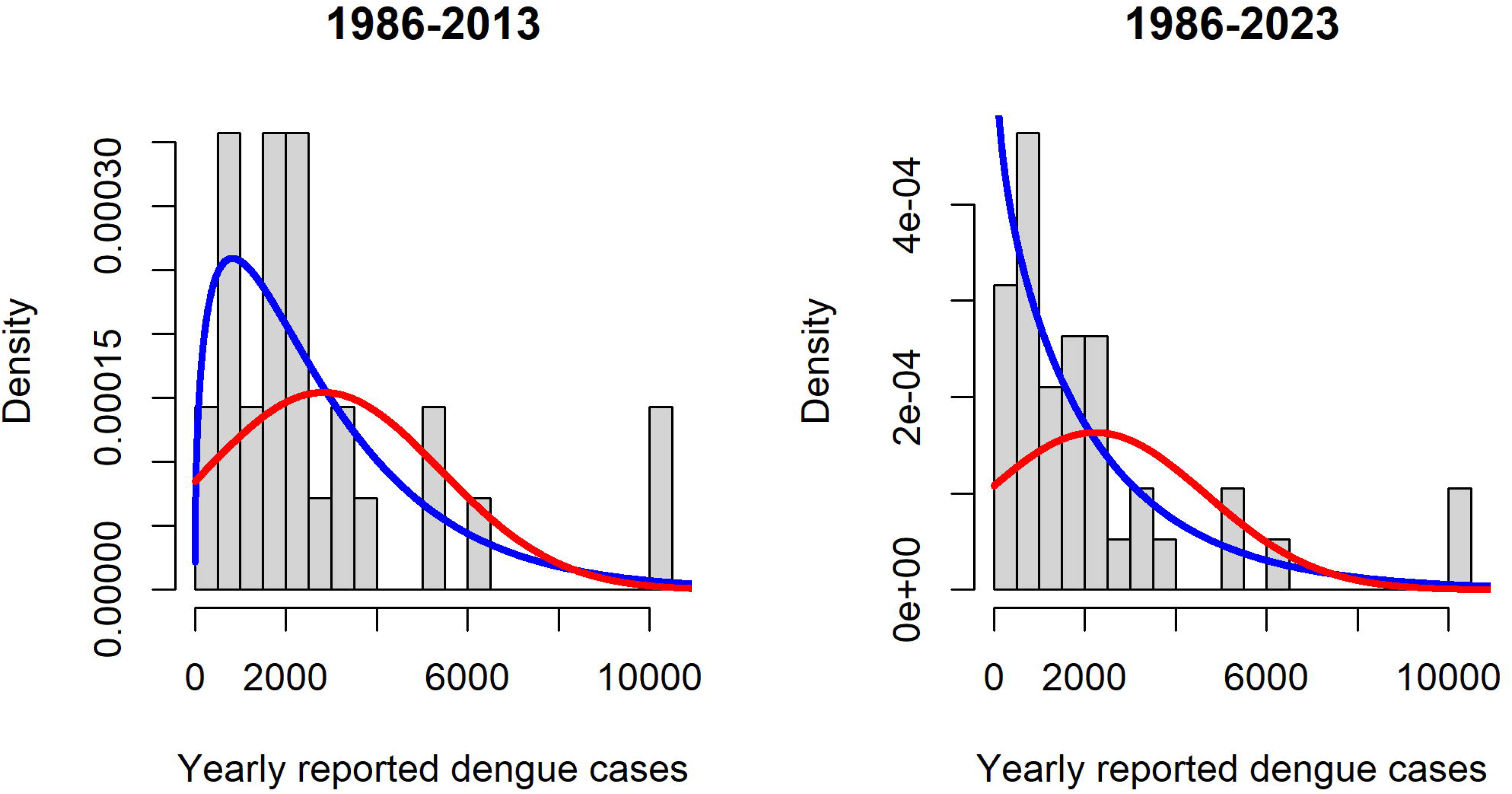

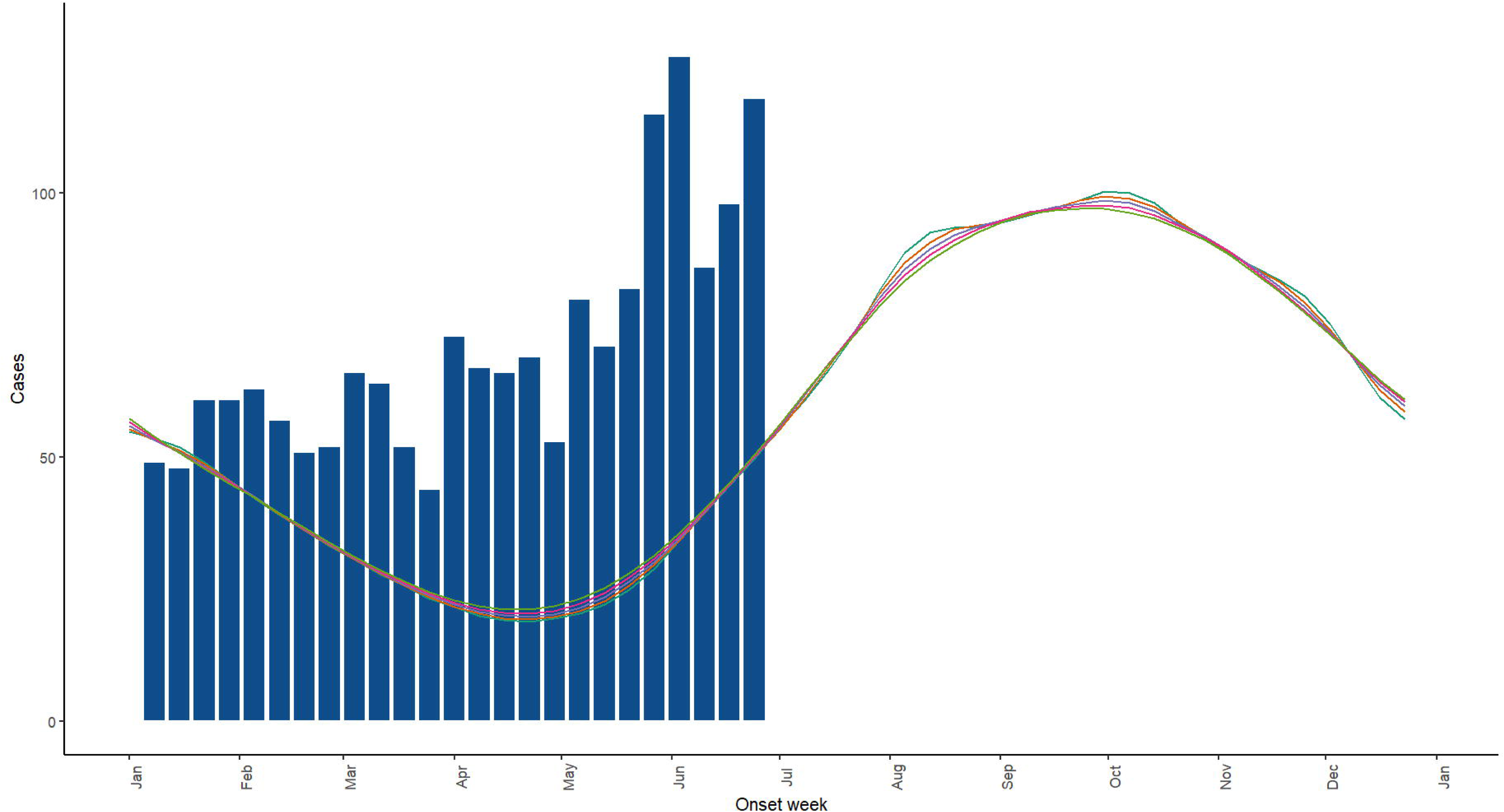

On the other hand, DENV cases began to increase in Puerto Rico at the beginning of 2024. The numbers exceeded the historical threshold in February 2024, a period when cases are generally low and were expected to be decreasing (Fig 4B). The dengue alert threshold, combined with a review of years when cases similarly exceeded the threshold early in the year (2009, 2010, 2013), supported the decision to declare an epidemic in Puerto Rico. Cases remained above the threshold starting in February and correctly predicted in the first dengue epidemic in over a decade.

## Discussion

Epidemic alert thresholds serve as critical tools for public health decision-making for preparedness and response to combat dengue epidemics. A well-defined and validated threshold provides an early yet reliable indication of an impending epidemic, allowing for timely public messaging, resource allocation planning, faster implementation of interventions like vector control, and risk communication strategies. Earlier response and interventions can help mitigate health risks and reduce healthcare costs or economic losses associated with epidemics.

The method presented here for classifying epidemics offers several advantages over traditional approaches used for epidemic detection. These traditional approaches use various tools, each with its own strengths and limitations. Early warning systems based on exceeding thresholds, for example, use predetermined thresholds based on specific indicators (e.g., case numbers) to trigger alerts (17). While simple and interpretable, these systems often rely on static thresholds that might not adapt to changing trends or skewed data distributions. Statistical anomaly detection, another approach, uses statistical techniques to identify unusual patterns in surveillance data that might signal an outbreak (18, 19). However, these methods can be sensitive to random fluctuations and might struggle to differentiate between true outbreaks and seasonal variations, potentially leading to missed detections or false alarms. Our approach acknowledges the skewed nature of dengue case counts and employs the negative binomial distribution, a more suitable statistical model for such data compared to methods assuming normality. Additionally, our method leverages all available historical data, strengthening the model’s robustness as more information is incorporated each year. This approach integrates outliers, including past epidemics, as valuable data points for building more informed thresholds. By using a negative binomial regression model, our method allows model-estimated medians and quantiles to naturally incorporate these outliers, leading to more comprehensive thresholds for epidemic detection.

Similar to the two standard deviation threshold used in most endemic channels to asses annual variability (9), our method requires choosing a percentile value to define the epidemic threshold. Two key factors can guide this choice, both related to the impact of large epidemics that would warrant additional response actions. First, historical data on epidemic occurrence in a specific region can inform the threshold selection. For instance, if major dengue epidemics in Puerto Rico typically occur every four or five years, thresholds set at the 75th or 80^th^ percentile might be appropriate. These higher percentiles capture a substantial increase in cases compared to typical years, potentially signaling an emerging epidemic. Second, we can gain valuable insights by fitting the thresholds retrospectively to historical data. By comparing these thresholds with known epidemic years (established by public health authorities), we can determine which percentile level aligns most effectively with past epidemic detections. This approach ensures the threshold accurately identifies periods requiring a heightened public health response.

While the negative binomial model offers a relatively straightforward approach to fitting epidemic alert thresholds, several limitations require consideration. First, the model relies on several years of consistent historical data to ensure robust model fitting. This can be a challenge in settings with limited or inconsistent surveillance data (20). Second, the use of all historical data assumes that it is representative of current patterns. If there is a reason to believe that trends have changed, the historical data could be restricted to exclude certain years. Additionally, the effectiveness of this method hinges on the availability of timely case reports for comparison with the thresholds. During outbreaks, reporting delays can be exacerbated due to overwhelmed surveillance systems (21). Nowcasting methods that estimate unreported cases and validated forecasts that assess the likelihood of exceeding the threshold in the near future could help mitigate these delays, as demonstrated for mpox and COVID-19 (22). Furthermore, the method can be less effective when dealing with extended periods of low or zero weekly case counts. This could occur during periods of minimal transmission or in settings with small populations. We can improve the model’s performance in sparse data scenarios by incorporating techniques like temporal bootstrapping or sparse approximate inference for spatiotemporal point processes (23). Future work could also explore more complex variations of the thresholds model, for example differentiating between higher and lower dengue case incidence years or adding additional predictors to the regression model.

Finally, while the thresholds presented here use data on probable and confirmed dengue cases, laboratory confirmation might not be readily available in all locations. However, the model can also use suspected case data for threshold development. In Puerto Rico, suspected case data were used before the introduction of chikungunya and Zika viruses, which share symptoms with dengue. The emergence of these viruses led to a rise in “suspected” dengue cases during the chikungunya outbreak in 2014 and the Zika outbreak in 2016, likely reflecting these other infections. Using confirmed and probable data addressed this issue. While the model can use suspected case data in settings with limited laboratory resources, this approach may include false positives, potentially leading to less accurate thresholds. In Puerto Rico, with its robust dengue surveillance system and routine testing for chikungunya and Zika, confirmed and probable cases provided the most reliable data for threshold development. The thresholds shown illustrate Puerto Rico as an example, but the thresholds are easily adaptable to other settings with different data types and patterns.

This evidence-based approach provided valuable real-time data to support communication with media and public partners, contextualizing the increase in dengue cases within historical norms in both 2022 and 2024. In 2024, this early identification of an epidemic via these epidemic alert thresholds allowed public health officials to respond quickly and appropriately before cases began to increase sharply above expected levels.

The utility of this tool lies in its real-time capability to identify and classify an epidemic alert. In Puerto Rico, the weekly historical median and epidemic alert thresholds are established annually in January for the upcoming year. Each week, as reported cases become available, the number of cases is compared to the weekly historical median and epidemic threshold to assess the current status of DENV transmission. If case counts exceed the threshold for two consecutive weeks, an epidemic alert is declared. This evidence-based, timely, and straightforward comparison and classification provides an ideal tool for guiding responses to emerging epidemics before high levels of DENV transmission are apparent. However, to maximize planning and response efforts, this method should be used in combination with other strategies and considered within the broader epidemiologic context. Recognizing high levels of DENV transmission is an important step, but it is also crucial to examine other criteria such as geographic regions and subpopulations where high levels are occurring. Using and evaluating these thresholds in conjunction with other tools, and as part of a larger discussion with key partners, can help support transparency and build trust between public health agencies and community members.

This threshold tool serves as an early warning system for dengue outbreaks, offering a crucial component within the broader public health surveillance and response framework. It can be combined with other tools for earlier identification and detection, such as integrating forecasts with thresholds or using methods with higher sensitivity and lower specificity. These methods can provide early indicators of increasing incidence. Although we have described these threshold methods in the context of dengue in Puerto Rico, they are adaptable to other geographical scales, locations, and diseases.

## Data Availability

All data produced in the present work are included in the linked GitHub repository in the manuscript.

https://github.com/maile-thayer/epidemic_thresholds

## References

1. U.S. Department of Health and Human Services. Principles of Epidemiology in Public Health Practice. Third Edition ed. Atlanta: Centers for Disease Control and Prevention,; 2012.

2. Shepard DS, Undurraga EA, Halasa YA, Stanaway JD. The global economic burden of dengue: a systematic analysis. The Lancet Infectious diseases. 2016 Apr 15.

3. Stanaway JD, Shepard DS, Undurraga EA, Halasa YA, Coffeng LE, Brady OJ, et al. The global burden of dengue: an analysis from the Global Burden of Disease Study 2013. The Lancet Infectious diseases. 2016 Feb 10.

4. Messina JP, Brady OJ, Golding N, Kraemer MUG, Wint GRW, Ray SE, et al. The current and future global distribution and population at risk of dengue. Nature Microbiology. 2019;4(9):1508–15.

5. Méndez-Lázaro P, Muller-Karger F, Otis D, Mccarthy M, Peña-Orellana M. Assessing Climate Variability Effects on Dengue Incidence in San Juan, Puerto Rico. International Journal of Environmental Research and Public Health. 2014 2014-09-11;11(9):9409–28.

6. Noyd DS, TM. Recent Advances in Dengue: Relevance to Puerto Rico. P R Health Sci. 2015;34(2):65–70.

7. Sharp TM, Hunsperger E, Muñoz-Jordán JL, Margolis HS, Tomashek KM. Sequential Episodes of Dengue—Puerto Rico, 2005–2010. The American Journal of Tropical Medicine and Hygiene. 2014;91(2):235–9.

8. Centers for Disease Control and Prevention. Informe Semanal de Vigilancia del Dengue: 1 al 7 de enero de 2014; 2014.

9. Bortman M. Elaboración de corredores o canales endémicos mediante planillas de cálculo. Rev Panam Salud Publica/Pan Am J Public Health. 1999;5(1).

10. Brady OJS, David L.; Scott, Thomas W.; Hay, Simon I. Dengue disease outbreak definitions are implicitly variable. Epidemics. 2015;11:92–102.

11. Hernández MA, Diana; Arce, Stephania; Benavides, Allan; Tejada, Paola Andrea; Ramírez, Sindy Vanessa; Cubides, Ángela. Metodología para la elaboración de canales endémicos y tendencia de la notificación del dengue, Valle del Cauca, Colombia, 2009-2013. Biomédica. 2016;36:98–107.

12. Badurdeen SBV, David; Farrar, Jeremy; Gozzer, Ernesto; Kroeger, Axel; Kuswara, Novia; Ranzinger, Silvia Runge; Tinh, Hien Tran; Leite, Priscila; Mahendradhata, Yodi; Skewes, Ronald; Verrall, Ayesha. Sharing experiences: towards an evidence based model of dengue surveillance and outbreak response in Latin America and Asia. BMC Public Health. 2013;13:607.

13. World Health Organization T. Operational guide using the web-based dashboard Early Warning and Response System (EWARS) for dengue outbreaks. Geneva; 2020.

14. R Core Team. R: A language and environment for statistical computing. Vienna, Austria: R Foundation for Statistical Computing; 2021.

15. R Core Team. The R Stats Package. 4.4.0 ed.

16. Venables WR, BD. Modern Applied Statistics with S. Fourth ed. New York: Springer; 2002.

17. Lowe R, Bailey TC, Stephenson DB, Graham RJ, Coelho CA, Carvalho MS, Barcellos C. Spatio-temporal modelling of climate-sensitive disease risk: Towards an early warning system for dengue in Brazil. Computers & Geosciences. 2011;37(3):371–81.

18. Eze PU, Geard N, Mueller I, Chades I. Anomaly Detection in Endemic Disease Surveillance Data Using Machine Learning Techniques. Healthcare (Basel). 2023 Jun 30;11(13).

19. Chen H, Zeng D, Yan P, Chen H, Zeng D, Yan P. Data analysis and outbreak detection. Infectious Disease Informatics: Syndromic Surveillance for Public Health and BioDefense. 2010:49–72.

20. Shadbolt N, Brett A, Chen M, Marion G, McKendrick IJ, Panovska-Griffiths J, et al. The challenges of data in future pandemics. Epidemics. 2022 Sep;40:100612.

21. McGough SF, Johansson MA, Lipsitch M, Menzies NA. Nowcasting by Bayesian Smoothing: A flexible, generalizable model for real-time epidemic tracking. PLOS Computational Biology. Apr 6, 2020;16(4).

22. Charniga K, Madewell ZJ, Masters NB, Asher J, Nakazawa Y, Spicknall IH. Nowcasting and forecasting the 2022 U.S. mpox outbreak: Support for public health decision making and lessons learned. Epidemics. 2024 Jun;47:100755.

23. Cseke B, Zammit-Mangion A, Heskes T, Sanguinetti G. Sparse Approximate Inference for Spatio-Temporal Point Process Models. Journal of the American Statistical Association. 2016 2016/10/01;111(516):1746–63.

